# Malaria diagnosis in rural healthcare facilities and treatment-seeking behavior in malaria endemic settings in western Kenya

**DOI:** 10.1101/2023.01.05.23284237

**Authors:** Collince J. Omondi, David Odongo, Wilfred O. Otambo, Kevin O. Ochwedo, Antony Otieno, Ming-Chieh Lee, James W. Kazura, Andrew K. Githeko, Guiyun Yan

## Abstract

Accurate diagnosis and timely treatment are central requirements for effective malaria management in communities. However, in resource-constrained settings, healthcare facilities are likely to be few, inaccessible, and ill-equipped with frequent drug or rapid diagnostic test kit (RDT) shortages. This may jeopardize much-needed quality care for patients and may have an impact on treatment-seeking behavior among the local population. The study’s goal is to determine treatment-seeking behavior, malaria diagnosis and treatment, and likely treatment-seeking determinants in the local population. Passive case detection, which targeted all patients with suspected malaria cases, was conducted in ten public healthcare facilities over a three-month period. Monthly malaria cases, methods of diagnosis and antimalarial drug availability were assessed. A household-based survey was also carried out. Structured questionnaires were used to collect data from household heads. Malaria knowledge, treatment seeking behavior, and predictors of malaria treatment-seeking were all determined. Three of the seven dispensaries lacked a laboratory to conduct microscopy-based diagnosis. These three dispensaries also experienced frequent RDT stock-outs, which resulted in a clinical diagnosis of malaria. The majority of local residents with fever (50.3%) purchased antimalarial drugs from a chemist. About 37% of fever patients sought treatment at healthcare facility while the remaining 12.7% did nothing. In irrigated areas, 45.5% (46/64) of fever patients sought treatment at healthcare facilities, compared to 25% (18/64) in non-irrigated areas (p = 0.009). Most children aged below 5 who had fever (77.7%) were taken to healthcare facility for treatment compared to 31.4% of older children or 20.9% of adults (0.0001). Predictors of treatment seeking included access to healthcare facility (OR = 16.23, 95% CI: 2.74-96.12), and ability to pay hospital bill (OR = 10.6, 95% CI: 1.97-57). Other factors that influenced health-seeking behavior included the severity of symptoms, the age of the fever patient and knowledge of malaria symptoms.

## Introduction

Malaria is still a major public health challenge in sub-Saharan Africa, with young children suffering the highest morbidity and mortality rates [3, 22, 58]. The debilitating nature of the disease is exacerbated by the overlap of malaria symptoms with those of other diseases [9, 21, 46]. As a result, accurate diagnosis combined with timely medication remains the primary life-saving intervention and means of reducing transmission [3, 55] Previously, the World Health Organization recommended treating any febrile patient with antimalarial drugs in the absence of microscopy or malaria diagnostic test kits [56]. However, this was later revised to emphasize parasite-based diagnosis in all ages. The widespread adoption of malaria diagnostic test kits (RDT) in healthcare facilities, as well as a decrease in febrile illness caused by malaria, prompted the revision [12, 56]. In addition, the emergence of drug-resistant malaria parasites due to drug overuse, possibly led to the implementation of the ‘test and treat’ strategy [19, 43, 57]. Microscopy and RDT kits were the two diagnostic tools widely used in healthcare facilities. Malaria RDT kits were relatively easy to use in malaria diagnosis [35]. This made parasitological diagnosis of malaria possible, especially in resource-poor healthcare facilities where microscopy diagnosis was a challenge [11].

In an effort to streamline with WHO recommendation, the Kenyan ministry of health endorsed the test and treat policy in all patients presenting with fever [32]. This ensures that only malaria-confirmed cases receive antimalarial treatment, as well as proper management of fever patients suffering from non-malarial illnesses [32]. Most importantly, it is prudent to minimize indiscriminate antimalarial prescription [61, 62]. However, treatment of suspected malaria based on clinical diagnosis is increasingly becoming common in most resource-poor African settings [4, 39, 48]. This situation has been aggravated by a number of factors, including ill-equipped healthcare facilities [26] and self-treatment among populations.

Significant reductions in parasite prevalence have been reported in Homa Bay county, where malaria infection was previously always above 20% [7, 31, 42]. Accurate diagnosis and proper treatment-seeking culture are critical for maintaining such gains and possibly reaching malaria-free Kenya, as envisioned in Kenya’s malaria strategy 2019-2023 [34]. However, studies investigating malaria diagnosis practice at the healthcare facilities and treatment-seeking around this region are limited. The study therefore seeks to investigate the probable challenges faced by healthcare facilities with regard to diagnosis and treatment as well as treatment-seeking behavior among the local population. The findings of the study will further guide the ministry of health and other stakeholders determine the best way to manage malaria disease.

## Methods

### Study site

The study was conducted in two sub-counties (Rachuonyo south and Rangwe) of Homa Bay county, Kenya. The County is situated in the southern part of former Nyanza Province and lies between latitude 0.15°S and 0·52° S and between longitude 34° E and 35° E. The government of Kenya initiated an irrigation scheme in parts of Homa Bay county to boost food production and household income [36]. The study targeted households within the irrigated (Oluch-Kimira irrigation scheme) and non-irrigated areas (5-10 km away from irrigation scheme). The study area experiences semi-arid climatic conditions where daily temperatures range between 26°C and 34°C during cold months (April and November) and hot months (January to March) respectively. It has two rainy seasons; long rains (April-June) and short rains (September-November) with rainfall ranging between 250mm and 1200mm annually.

The major economic activity is subsistence farming where crops such as rice, maize beans cabbages and variety of fruits are grown. Other economic activities practiced in the area include small scale businesses, fishing, while some are government employees. Malaria transmission within the county is perennial with a parasite prevalence of 27%. [30] The primary vector control strategy in this county has been long lasting insecticide treated nets, but in February 2018 indoor residual spray program using Actellic insecticide was implemented [45]. Significant progress in malaria reduction has been reported since the implementation of IRS [42].

### Study design and measures

The study was a cross-sectional household and healthcare facility-based survey. Healthcare facility survey was carried out between October 2019 and December 2019 coinciding with the short rainy season. Healthcare facilities with a minimum catchment population of 3000 people were selected. As a result, ten government healthcare facilities were identified and recruited into the study due to low cost or free services offered there. Table 1 further describes the total number of villages, households and population per healthcare facility. According to the Kenya national guidelines for diagnosis, treatment, and control, all patients in all age groups with suspected malaria should undergo a parasite-based diagnosis before treatment [32]. Microscopy and malaria rapid diagnostic test kits (RDT) were some of the recommended malaria diagnostic tools particularly in resource-poor areas [32]. The study therefore aimed to determine the average monthly clinical malaria cases and to assess the available diagnostic techniques for suspected malaria cases. The study targeted all patients with suspected malaria attending the ten government-based healthcare facilities and agreed to sign informed consent or assent form (for minors aged <18 years old). Besides, the patient had to be a resident of the study area in order to participate. Clinical officer in charge of each healthcare facility provided information on Artemether Lumefantrine (AL) availability. AL is the recommended first-line drug for the treatment of uncomplicated *Plasmodium* infections in Kenya [32].

**Table 1.** List of the 10 healthcare facilities within the study area

| Name | level | Catchment population |  |  | Type of malaria diagnosis offered |
| --- | --- | --- | --- | --- | --- |
|  |  | No. of villages | No. of households | Total population |  |
| Nyarut | Dispensary | 18 | 792 | 3,774 | RDT, clinical |
| Kodula | Dispensary | 12 | 930 | 6,797 | RDT, clinical |
| Omboga | Dispensary | 25 | 2165 | 9,937 | RDT, clinical |
| Ngegu | Dispensary | 20 | 1614 | 7,760 | RDT, microscopy |
| Nyagoro | Health center | 34 | 3203 | 15,583 | RDT, microscopy |
| Nyalkinyi | Health center | 20 | 2147 | 10,730 | RDT, microscopy |
| Okiki Amayo | Dispensary | 10 | 477 | 3,142 | RDT, microscopy |
| Adiedo | Dispensary | 29 | 1727 | 9,234 | RDT, microscopy |
| Obwanda | Dispensary | 16 | 927 | 4,864 | RDT, microscopy |
| Kandiego | Sub-county hospital | 51 | 2809 | 15,338 | RDT, microscopy |

### Household survey

Household-based survey was conducted in the month of July 2020, near the end of long rains, when malaria transmission typically at its peak. Households were the study units in the selected villages. A total of 240 households were considered sufficient to answer study objectives and were randomly selected using a two-stage cluster sampling design as previously described [15]. First, the ten health facilities served a total of 208 villages in both irrigated and non-irrigated areas. There were 73 villages in irrigated area and 135 villages in non-irrigated areas. Based on the number of households in each village, 12 villages (6 from each region) were randomly selected. The second stage involved household selection. All households in the 12 identified clusters (villages) were assigned unique numbers by the study team. Thereafter, 20 households in each of the 12 clusters, were chosen at random.

Trained interviewers distributed structured questionnaires to the household heads. All 240 household heads responded to questions regarding knowledge, attitude and practice towards malaria. However, to determine the treatment seeking behavior, only households that had reported cases of fever accompanied by headache or joint pains or both in the two weeks preceding data collection, were interviewed. The household head provided information on the family members who had fever (age, gender and severity of fever). The household head also answered questions about treatment-seeking behavior on behalf of febrile patients, bed net ownership, and use. The integrity of nets was physically examined. Health-seeking behavior was the primary outcome of the study. The operational definition of treatment-seeking behavior was considered to be seeking treatment at health facility or self-medication using antimalarial drugs bought from a chemist. The proportion of participants who did nothing to treat their fever was also recorded. We also enquired whether antimalarial drugs were available in healthcare facilities for fever patients seeking treatment, or if they were referred to a chemist.

Potential covariates to treatment seeking behavior such as easy access to hospital, ability to pay hospital bills, the nature of symptoms (severe or mild), age group, whether the house was sprayed (IRS), knowledge of malaria symptoms, region of residence (irrigated or non-irrigated), household head level of education and use of a bed net were considered and included in a predictive model. The level of education for the household head was categorized into no education, primary education, secondary education and college education. Assessment of knowledge of malaria symptoms was based on the ability of the household head to link fever or headache to malaria.

### Data analysis

The data were coded, entered in Excel sheet, and analyzed using GMP Pro 16. Demographic characteristic of participants, knowledge, attitude and practice towards malaria were summarized using descriptive statistics. Differences in proportions were compared using either Fisher’s exact test or Chi-square test. To assess treatment-seeking behavior, we first used descriptive analysis to determine the difference among those who sought treatment at health facility versus those who used antimalarial drugs bought from the chemist or those who did nothing to manage the fever. We then used multiple logistic regression to identify the factors that influence treatment-seeking behavior.

### Ethical consideration

The current study was part of a larger survey on “Environmental Modifications in sub-Saharan Africa: Changing Epidemiology, Transmission and Pathogenesis of *Plasmodium falciparum* and *P. vivax* malaria” whose ethical review and approval (MSU/DRPI/MUERC/00456/17) was obtained from Maseno University (Kenya) Ethics and Research Committees and University of California, USA. Written informed consent was obtained from the parent/guardian of each participant under 18 years of age.

## Results

### Socio-demographic characteristics of study participants in household survey

We visited 240 households with a total population of 1,142 people (574 from irrigated and 568 from non-irrigated areas). Children under the age of 5 constituted 20.2% (231/1142), followed by children aged 5 to 14 years (32.9%, 376/1142), and those aged 15 years and older constituted 46.8 (535/1142). The mean household size was 4.8 people, with a range of 1-11individuals per household. The majority of the respondents were females (76.3%, n =183) as indicated in table 2. Most of the respondents (58.8%, n = 141) had a primary education, while the least (4.2%, n = 10) had a college education. The overall bed net ownership by study participants was 93.3% (224/240). Irrigated area had significantly higher bed net ownership of 98.3% compared to that of non-irrigated area (88.3%) (Fisher’s exact test = 0.003). The proportion of households having 1 LLIN for two people was 37.5% (45/120) in irrigated compared to 24.2% (29/120) in non-irrigated area (χ^2^= 5.0, df = 1, p = 0.03). Bed net usage survey indicated that in most households (83.3% in irrigated and 73.7% in non-irrigated area), all family members used bed net. Although bed net ownership and usage was above average in the study area, the majority of the households (59.2% in irrigated and 57.5% in non-irrigated areas) had nets which were torn. Indoor residual spray coverage was at 66.7% (80/120) in irrigated and 69.2% (83/120) in non-irrigated areas.

**Table 2.** Demographic information of the households visited and the household heads

| Variables | Irrigated (n = 120)<br>n (%) | Non-irrigated (n = 120)<br>n (%) | p-value |
| --- | --- | --- | --- |
| Sex of respondent |  |  |  |
| Male | 26 (21.7) | 31 (21.8) | 0.5 |
| Female | 94 (78.3) | 89 (74.2) |  |
| Education |  |  |  |
| No education | 14 (11.7) | 10 (8.3) | 0.5 |
| Primary | 61 (50.8) | 80 (66.7) | 0.02 |
| Secondary | 36 (30.0) | 29 (24.2) | 0.38 |
| College | 9 (7.5) | 1 (0.8) | 0.019 |
| Bed net ownership | 118 (98.3) | 106 (88.3) | 0.003 |
| All family members sleep under bed net? |  |  |  |
| Yes | 100 (83.3) | 88 (73.7) | 0.08 |
| No | 20 (16.7) | 32 (26.7) |  |
| 1 LLIN for 2 people | 45 (37.5) | 29 (24.2) | 0.03 |
| IRS in the last 6 months? |  |  |  |
| Yes | 80 (66.7) | 83 (69.2) | 0.8 |
| No | 40 (33.3) | 37 (55.8) |  |

### Monthly clinical malaria and diagnosis routine at the healthcare facilities

The majority of public healthcare facilities 70% (7/10) in the study area are dispensaries which are headed by clinical officers and provide only outpatient services. Health centers and a hospital which are headed by at least one doctor and provide inpatient services accounts for 20% (2/10) and 10% (1/10) respectively. A total of 1264 suspected malaria cases in the 10 healthcare facilities were recorded during a period of 3 months. Out of the 1264 cases, 76.7% (972/1264) were diagnosed by microscopy, 4.2% (53/1264) by RDT kits while 18.9% (239/1264) were clinically diagnosed. The overall clinical malaria cases were 19.7% (65/330), 18.2% (109/600), and 21% (70/334) during October, November and December respectively as illustrated in table 3

**Table 3.** Monthly suspected and confirmed malaria cases from the 10 selected health facilities within the study area

|  | October 2019 |  | November 2019 |  | December 2019 |  |
| --- | --- | --- | --- | --- | --- | --- |
| Type of diagnosis | Suspected malaria | Positive n (%) | Suspected malaria | Positive n (%) | Suspected malaria | Positive n (%) |
| Microscopy | 241 | 39 (16.2) | 466 | 70 (15.0) | 265 | 47 (17.7) |
| RDT | 27 | 3 (11.1) | 21 | 5 (23.8) | 5 | 0 (0) |
| Clinical | 62 | 23 (38.0) | 113 | 34 (30.1) | 64 | 23 (35.6) |

During the three months of the study, three dispensaries (Nyarut, Omboga, and Kodula), did not have RDT kits and thus relied on clinical symptoms to confirm suspected malaria cases. The number of cases confirmed by clinical diagnosis was always greater than 30% during the 3 months of the study. Microscopy confirmed cases, however, accounted for less than 20% of all cases during the same time period. For example, during the month of October 2019, the proportion of suspected malaria which were confirmed by microscopy was 16.2% (39/241) compared 38% (23/62) clinically confirmed cases (p = 0.0005). During the month of November 2019, the proportion of microscopy confirmed cases was 15% (70/466) compared to 30.1% (34/113) cases confirmed clinically (p = 0.0003). Similarly, during the month of December 2019, cases confirmed by microscopy were 17.7% (47/265) while those confirmed clinically were 35.6% (23/64) (p = 0.0025) (Table 4).

**Table 4.** Comparison between Slide positivity rate (SPR) and clinically confirmed cases at health care systems

|  | October 2019 |  | November 2019 |  | December 2019 |  |
| --- | --- | --- | --- | --- | --- | --- |
|  | Confirmed cases n (%) | p-value | Confirmed cases n (%) | p-value | Confirmed cases n (%) | p-value |
| SPR | 39 (16.2) | 0.0005 | 70 (15.0) | 0.0003 | 47 (17.7) | 0.0025 |
| Clinical | 23 (38.0) |  | 34 (30.1) |  | 23 (35.6) |  |

### Knowledge, attitude and practice of household heads towards malaria

Table 5 summarizes the knowledge, attitude and practice of respondents towards malaria. The majority of respondents (77%), indicated that malaria is transmitted by the bite of any mosquito. Other means of malaria transmission mentioned included the bite of infected mosquito (11.3%), and contaminated water (5.4%). About 8.3% however, indicated that they did not know how malaria is transmitted. With regards to malaria symptoms, the following symptoms were identified by the village residents; fever 177 (73.8%), headache 171 (71.3%), joint pains 120 (50%) and vomiting 95 (39.6%). Children under the age of 5 years, were mentioned as the most vulnerable 174 (72.5%) group. Other vulnerable groups included, pregnant women 108 (45%), the elderly 50 (20.8%), everyone 63 (26.3%), and older children 4 (1.7%). The majority of participants 288 (95%) indicated that the use of treated bed-nets was the best way to control malaria. Other control methods mentioned included: mosquito coils/ repellants 47 (19.6%), indoor residual spray 44 (18.3%), chemoprophylaxis 32 (13.3%), and keeping food clean 22 (9.2%). The knowledge about mosquito breeding habitat was assessed and the findings indicate that 95% (228/240) of the study participants were aware that stagnant water is the most suitable breeding place of mosquitos. The other breeding place mentioned was bushes at 66.7% (166/240). Attitude towards malaria was also assessed and 95.4% (229/240) of the participants indicated that malaria was a serious disease while 4.6% (11/240) mentioned that malaria is a mild disease. Approximately 93.8% (225/240) indicated that it was very important to follow malaria treatment prescription given by the doctor, while 6.2% (15/240) indicated that it was not important.

**Table 5.** Knowledge, attitude, and practice towards malaria

| **Characteristics** | **Frequency (%)** |
| --- | --- |
| **How can malaria be transmitted (n= 240)** |  |
| Bite of any mosquito | 185 (77) |
| Bite of infected mosquito | 27 (11.3) |
| Contaminated water | 13 (5.4) |
| Do not know | 20 (8.3) |
| **Malaria symptoms known (n= 240)** |  |
| Fever | 177 (73.8) |
| Headache | 171 (71.3) |
| Joint pains | 120 (50) |
| Vomiting | 95 (39.6) |
| Chills | 19 (7.9) |
| Loss of appetite | 15 (6.3) |
| Common cold | 44 (18.3) |
| Do not know | 2 (0.8) |
| **Who are most vulnerable towards malaria (n = 240)** |  |
| Everyone | 63 (26.3) |
| Children below 5 years old | 174 (72.5) |
| Pregnant women | 108 (45) |
| Children above 5 years | 4 (1.7) |
| Adults | 50 (20.8) |
| Don't know | 8 (3.3) |
| **Malaria control methods (n = 240)** |  |
| Insecticide treated nets | 228 (95) |
| Indoor residual spray | 44 (18.3) |
| Mosquito coils/ repellants | 47 (19.6) |
| Window screen | 4 (1.7) |
| Chemoprophylaxis | 32 (13.3) |
| Avoid playing in the cold | 14 (5.8) |
| Do not know | 6 (2.5) |
| Keeping food clean | 22 (9.2) |
| **Breeding places of mosquito (n = 240)** |  |
| Stagnant water | 228 (95) |
| Bushes | 160 (66.7) |
| Do not know | 8 (3.3) |
| **How serious is malaria to human health (n = 240)** |  |
| Very serious | 229 (95.4) |
| Mild | 11 (4.6) |

### Treatment-seeking pattern among study participants

Treatment-seeking behavior among fever patients is described in table 4. Out of the 240 households visited, 44% (106/240) households reported to have had fever cases in the previous 2 weeks. The total number of people who experienced fever cases were 15.1% (173/1142). The irrigated areas had a significantly higher number of fever patients (17.6%, n = 101) compared to that of non-irrigated (12.8%, n = 72) (χ^2^ = 5.0, df = 1, p = 0.03). Among these 173 individuals, 37.0% (n = 64) sought treatment at the health facility, 50.3% (n = 87) used antimalarial drugs bought from the chemist while 12.7% (n = 22) did nothing to manage the fever. The proportion of individuals seeking treatment at a health facility for fever cases were more common in irrigated (45.5%, n = 46) than in non-irrigated areas (25%, n = 18) (χ^2^ = 6.8, df = 1, p = 0.009) as shown in table 6. Malaria was confirmed in 65.6% (42/64) of the febrile patients who sought treatment at healthcare facilities. Due to drug stock-outs in some healthcare facilities, 87.7% (30/42) of the malaria-confirmed patients were directed to purchase antimalarial drugs from a chemist. In non-irrigated areas, most fever patients who did not seek treatment at healthcare facilities, bought drugs at the local chemists. For instance, pharmacy-purchased medications were used by 59.7% (43/72) of fever patients in non-irrigated areas compared to 43.6% (44/101) of fever patients in irrigated (χ^2^ = 3.8, df = 1, p = 0.05). Similarly, the proportion of those who did nothing or waited to get better when they experienced fever was 10.9% in irrigated and 15.3% in non-irrigated areas (χ^2^ = 0.4, df = 1, p = 0.5).

**Table 6.**
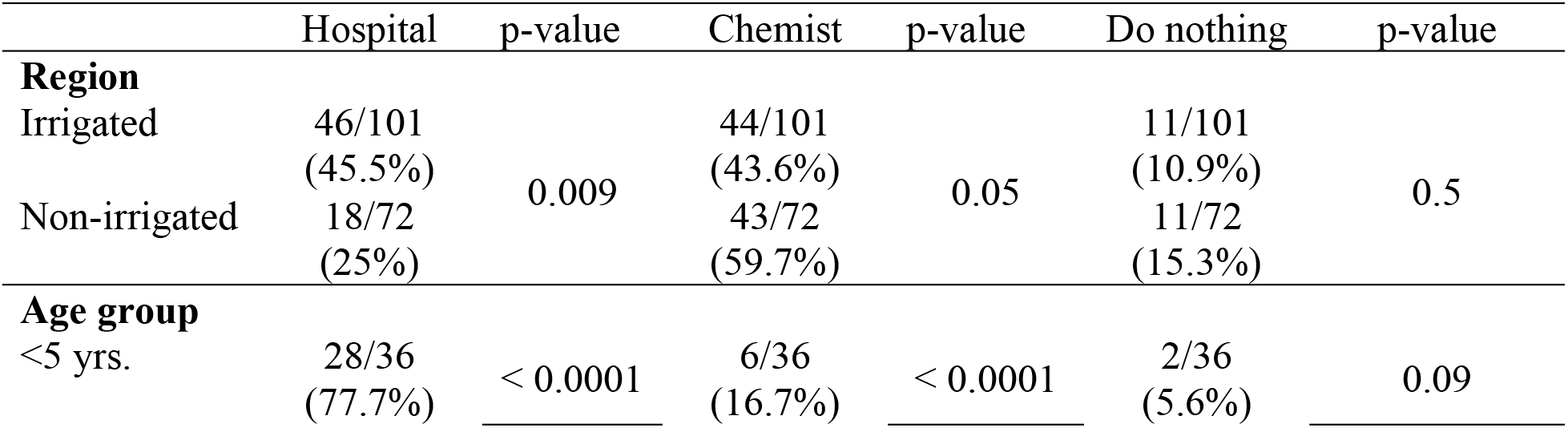
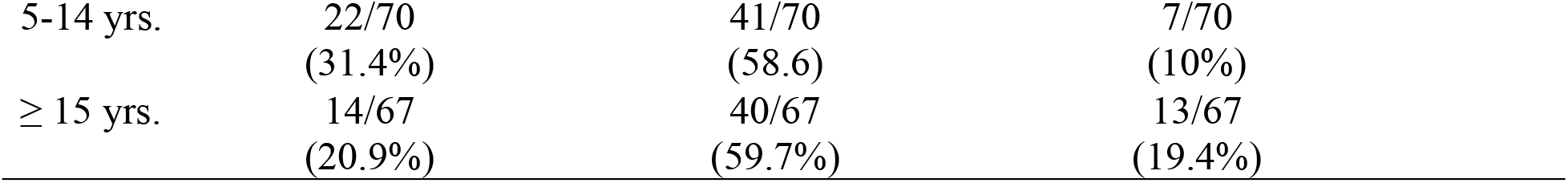
Malaria treatment seeking behavior (n = 173)

Treatment seeking behavior differed significantly among age groups. The majority of children aged < 5 years (77.7%) were taken to a health facility for treatment compared to 31.4% and 20.9% of children aged 5-14 years old and those aged ≥15 years respectively (χ^2^ = 37.07, df = 2, p < 0.0001). However, use of antimalarial drugs purchased from a chemist for fever-related cases was highest among adults (59.7%), followed by 58.6% of children aged 5-14, and lowest among children under the age of 5 (16.7%) (χ^2^ = 20.6, df = 2, p <0.0001). Approximately 5.6%, 10% and 19.4% of children under the age of 5, children aged 5-14 and those aged ≥15 years old, respectively, did not receive any form of treatment and instead their caregivers just waited for the fever to subside (χ^2^ = 4.8, df = 2, p = 0.09).

### Factors related to treatment-seeking

Fever patients were either treated at a health facility, used drugs bought from a chemist, or did nothing. Access to a health facility was one of the factors that influenced participants’ decision to seek treatment at a health facility. Those who had easy access to a hospital were more likely to seek treatment there (OR = 16.23, p = 0.002). than those who had a challenge accessing the health facility as indicated in table 7. Additional hospital costs, such as consultation fees or the cost of laboratory tests, influenced whether or not the patient sought treatment at the hospital. For example, those who indicated that paying medical bills was not a challenge to them, were more likely to seek treatment at the health facility (OR = 10.6, p = 0.006) than those who could not afford such costs. Those who experienced severe symptoms in the previous 2 weeks were more likely to seek treatment at the hospital (OR = 7.5, p = 0.037) than those who had mild symptoms. Care givers of under 5-year-old fever patients were also more likely to take their children to health facility for treatment (OR = 17.8, p = 0.008) than those caregivers of older children and adults.

**Table 7.** Nominal logistic fit for treatment seeking behavior

| **Hospital treatment** |  |  |  |
| --- | --- | --- | --- |
| Variable | Level | Odds ratio<br>95% CI | p-value |
| Easy access to hospital | Yes | 16.23 (2.74-96.12) | 0.002 |
|  | No | 1.00 | - |
| Ability to pay hospital bills | Yes | 10.60 (1.97-57.1) | 0.006 |
|  | No | 1.00 | - |
| Severe symptoms | Yes | 7.55 (1.12-50.80) | 0.037 |
|  | No | 1.00 | - |
| Age group | < 5yrs | 17.81 (2.12-149.7) | 0.008 |
|  | ≥5 yrs. | 1.00 | - |
| **Buy drugs at the chemist** |  |  |  |
| House sprayed (IRS) | Yes | 1.00 | - |
|  | No | 3.14 (1.15-8.57) | 0.025 |
| Mild symptoms | Yes | 12.09 (2.70-54.24) | 0.001 |
|  | No | 1.00 | - |
| Knowledge of malaria symptoms | Yes | 5.72 (1.87-17.49) | 0.002 |
|  | No | 1.00 | - |
| Age group | <5yrs | 1.00 | - |
|  | ≥5yrs | 4.82 (1.19-19.61) | 0.03 |

1 = Reference value

With regard to use of antimalarial drugs bought at the chemist, family members from houses which were not sprayed (IRS), were more likely to buy antimalarial drugs from the chemist (OR = 3.14, p = 0.025) than those from sprayed households. Study participants who experienced mild fever or headache, were more likely to buy antimalarial drugs from the chemist (OR = 10.09, p = 0.001) than those who experienced severe fever or headache. Knowledge of malaria symptoms also played key role towards buying of antimalarial. Those who believed that fever or headache was due to malaria, were more likely to buy drugs at the chemist (OR = 5.72, p = 0.002) than to go to hospital for treatment. Caregivers of older children or adults were 4 times more likely to buy antimalarial drugs at the chemist (OR = 4.82, p = 0.03) than seeking treatment at the hospital.

This study assessed the rural population’s knowledge, attitude, and practice regarding malaria, as well as treatment-seeking behavior and predictors of treatment-seeking in a resource-poor setting where malaria is endemic. Malaria knowledge and proper treatment seeking behavior are critical components of malaria control or elimination strategies. [17]. As a result, the findings of this study may have a positive impact on the current malaria control strategies, which have been intensified in order to reduce disease rates in the study area. Our findings revealed a community that is well-versed in malaria transmission, symptoms, and control methods. In most malaria endemic regions, the majority of residents are aware about malaria transmission and symptoms [2, 40].

Despite widespread knowledge of malaria control methods, study’s findings revealed that only a small percentage of households met the requirement of “one LLIN for every two people”. A minimum coverage of 60% is required for LLINs to provide optimal protection [24]. The present study’s 30.8% universal coverage ownership of LLINs was lower than the coverage (45%) recorded right after the completion of the 2017 mass distribution of nets. [33]. The survey was done three years after the previous mass distribution, thus most nets must have worn out and stopped being used, lowering the proportion of persons who had nets. Furthermore, the majority of nets surveyed had holes, decreasing nets’ ability to offer effective protection.

The findings further indicated that the majority of participants consider malaria to be a serious disease, acknowledge stagnant water as the suitable breeding place for mosquitoes, and are aware of the groups of people who are most vulnerable to malaria. This high level of knowledge in malaria disease may be attributed to the community’s location in a malaria endemic region where the disease is common, and also the fact that the majority of the respondents (90%) had at least a primary education. This is comparable to other studies conducted in southeastern Iran [2], in Mozambique [13], Northwestern Ethiopia [14], and Bangladesh [49].

According to the study findings, 44% of the total households surveyed had at least one fever patient. More than a half of these febrile cases were treated with antimalarial drugs obtained from a local chemist. This is similar to studies in in Indonesia [16], Ghana [5], India [50], Myanmar [53], and Bangladesh [2] where majority of febrile patients reportedly preferred self-medication over hospital-based treatment. In the current study, key predictors of self-treatment included knowledge of malaria symptoms, severity of symptoms, and age of febrile patients. The study area being a malaria endemic region, the majority of people are frequently exposed to *Plasmodium* infections. As a result, many participants who were familiar with malaria symptoms, were more likely to delay or fail to seek treatment at a health facility when fever developed. This finding is comparable to studies conducted in Ethiopia [18, 52]. However, the current study findings, contradicted previous research that found that knowledge about malaria increased the likelihood of early treatment-seeking in a health facility [29, 59]. The study further demonstrated that patients with mild symptoms were more likely to buy drugs from a chemist or do nothing. In all regions where malaria transmission is perennial, most people, particularly older children and adults, may be asymptomatic or exhibit mild clinical symptoms of malaria due to acquired immunity to the disease [6, 28, 54]. Therefore, people with mild malaria symptoms may delay or avoid seeking treatment at the hospital as previously reported [20].

Access to healthcare facilities, ability to pay hospital bills, severity of symptoms, and age of fever patients were factors that predicted treatment-seeking at healthcare facilities. The ease of access to healthcare facilities has been reported in previous studies to encourage treatment-seeking at healthcare facilities [16, 25, 37, 60]. Easily accessible health facilities may not require transportation or may require only a small amount of money; as a result, the majority of people find it easy to access them. Similarly, unavailability of money to pay for hospital bills such as laboratory tests charges discouraged some participants to seek treatment at these public healthcare facilities. Financial constraints have been reported in previous studies to have a significant impact on treatment-seeking at healthcare facilities [50, 51].

The study’s findings also demonstrated that young children with fevers, were more likely to be taken to the healthcare facility by their caregivers than older children or adults. This could be due to the majority of respondents being aware that children under the age of five are more vulnerable to malaria than older children or adults, as previously reported [1, 8]. Individuals who are more susceptible to malaria are more likely to experience severe symptoms. In the current study, most participants indicated that malaria is a serious disease. This could be the most plausible explanation for why the majority of people with severe symptoms sought treatment at healthcare facilities. This is comparable to a study in Cameroon in which people could only visit healthcare facilities if they experienced severe symptoms [27].

According to survey findings, some dispensaries were resource constrained. Chronic challenges that plagued the day-to-day operations of most of these dispensaries included a lack of well-equipped laboratories, RDT kits stock-outs, and antimalarial drug shortages. These challenges if not addressed, may have an impact on treatment seeking behavior. For example, patients who are referred to some distant healthcare facilities for proper checkups may resort to buying drugs at local chemists due to a lack of funds for transportation. Similarly, drug shortages in healthcare facilities are likely to hamper effective treatment seeking as reported in previous studies [10]. For instance, in irrigated areas, the majority of fever patients sought treatment at healthcare facilities compared to fever patients in non-irrigated areas. This could be due to a combination of factors, including limited access to healthcare facilities and the types of services provided at those facilities. For example, in irrigated areas, the majority of people have easy access to healthcare, as opposed to non-irrigated areas. Secondly, as opposed to non-irrigated areas, most healthcare facilities in the irrigated areas had well-established laboratories for conducting quality malaria diagnosis. These two factors may influence the treatment seeking behavior of local residents [16, 17]. Another plausible explanation could be income disparities between irrigated and non-irrigated households. A recent study conducted by Omondi, Ochwedo (41) in the same study area, discovered that household income in irrigated areas was higher than in non-irrigated areas.

World Health Organization recommends that all suspected malaria cases be confirmed through microscopy or the use of rapid diagnostic test kits before antimalarial drugs are administered [57]. Particularly in malaria-endemic areas, where the majority of febrile illnesses may not be caused by malaria [12, 23]. Although we did not re-examine to confirm the accuracy of clinically diagnosed cases, it was apparent that dispensaries that relied on clinical diagnosis reported a significantly higher number of malaria cases than those that used microscopy. This raises concerns about over-diagnosis of malaria cases, which could lead to indiscriminate use of antimalarial drugs or non-treatment of other illnesses with similar symptoms to malaria [38, 44, 47].

## Conclusion

The study findings indicate that most people from this region have good knowledge about malaria. However, the majority of fever patients self-treat with drugs purchased from local chemists. Access to healthcare facilities, knowledge of malaria symptoms, age of febrile patients, severity of symptoms, and income were all likely factors influencing treatment seeking behavior. Antimalarial drugs and RDT kits stock outs were common challenges to some dispensaries. The Ministry of Health should consider launching a community-based awareness campaign to educate local population about the importance of seeking medical attention at a healthcare facility. The ministry of health should also consider improving access to healthcare facilities as well as quality services in terms of diagnosis and drug availability.

## Data Availability

The dataset used in this study is available from the corresponding author upon reasonable request.

## Acknowledgements

We are grateful to the household heads who allowed us to conduct this research, and we appreciate your patience in responding to our questionnaires. We also like to thank the healthcare facility management and staff for their assistance throughout the study, which made data collection a success. This research would not have been possible without the permission of the Homa Bay County Ministry of Health. Finally, we would like to thank the International Center for Malaria Research staff for their assistance during data collection.

## Authors contribution

Conceptualization of the study: Collince J. Omondi, David Odongo, James W. Kazura, Andrew K. Githeko, Guiyun Yan

Data collection, analysis and interpretation: Collince J. Omondi

Methodology: Collince J. Omondi, Kevin Ochwedo, Wilfred Otambo, Ming-Chieh Lee, Andrew K. Githeko

Writing first draft: Collince J. Omondi

Manuscript review: David Odongo, Antony Otieno, James W. Kazura, Andrew K. Githeko, Guiyun Yan

